# For clinical data extraction, QLoRA attains accuracy close to LoRA while requiring lower compute resources

**DOI:** 10.1101/2025.10.21.25338506

**Authors:** Prabin R. Shakya, Ayush Khaneja, Kavishwar B. Wagholikar

## Abstract

**Background:** Large language models (LLMs) can accurately extract structured data from free text, yet fine-tuning for specific clinical tasks is often compute- and memory-intensive. We examine whether Parameter-Efficient Fine-Tuning (PEFT)—updating only a small subset of weights in the LLM— preserves accuracy on quantized models while further reducing memory and graphical processing unit (GPU) requirements for hardware-limited teams.

**Methods:** We fine-tuned three Llama-3.1-8B-Instruct variants: (i) a non-quantized low-rank adaptation (LoRA) model and (ii–iii) quantized low-rank adaptation (QLoRA) models initialized from 8-bit and 4-bit quantized bases. We used the ELMTEX corpus of 60,000 clinical summaries extracted from PubMed Central, which included manual annotations for 15 categories. Models were evaluated with naïve and advanced prompting to extract data from the corpus for the 15 categories. Advanced prompting involved a detailed task description and three examples selected using similarity scores. Metrics included ROUGE and BERTScore for lexical/semantic alignment, and entity-level precision, recall, and F1 to assess clinical concept extraction.

**Results:** Fine-tuning consistently outperformed prompting alone. LoRA improved metrics by 10–20 points over the base model, while QLoRA improved by 8–14 points—only 2–4 points below LoRA. Quantization lowered the need for resources— LoRA required 4 GPUs, versus 3 (8-bit) and 2 (4-bit) for QLoRA. Compared with LoRA, 4-bit QLoRA used about two-thirds of the peak GPU RAM. However training of quantized models took 28–32% longer, likely due to dequantization overhead and less-mature library routines.

**Conclusion:** PEFT on quantized models preserves most of LoRA’s accuracy gains while substantially reducing GPU count and memory footprint, providing a practical path for accurate clinical information extraction in resource-constrained settings. This study was limited to a single architecture (Llama-3.1-8B) and use of clinical summaries that are less complex than routine clinical notes, which constrains generalizability of the results. Future work should test QLoRA across diverse architectures and sizes and on clinical corpora representative of real-world practice.

## Background

Large language models (LLMs) can extract structured information from free text with high reliability. ^1,2^ However, their broad adoption is constrained by the substantial compute and memory demands to fine-tune the models for specific clinical tasks. Parameter-Efficient Fine-Tuning (PEFT) addresses this challenge by updating only a small subset of weights, sharply reducing the computational footprint while preserving near–full-tuning performance. ^3^ Additional reduction in computational needs may come from quantizing the models before training, but with potential loss of accuracy. In this paper we specifically ask whether applying PEFT to quantized models retains the accuracy boosts seen with PEFT on full-precision models, while cutting memory footprint and Graphical Processing Unit (GPU) needs for clinical data extraction. If successful, this approach would enable accurate, task-specific LLM extractors to run on modest hardware—lowering costs and widening access to LLM-driven clinical informatics across diverse health systems.

Over the past several decades clinical NLP has evolved from ruled-based and ontology based approaches, machine learning and deep learning based approaches, and recently, large language models (LLMs) have be extensively researched on clinical text. ^2,4–9^ LLMs are essentially typically transformers-based deep neural networks that are trained on the large amount of textual data. In addition to their high performance accuracy, these models can be invoked to perform data extraction tasks with simple instructions, bypassing the need for extensive programming that was required with previous approaches. ^10–12^

Multitude of LLM approaches had been researched for the extraction like few-shot relation extraction, soft-tunning or prompt tuning, and fine-tunning. ^13,14^ Fine-tuning has been shown to substantially improve accuracy over other methods; however, it requires intensive computational resources making it prohibitive for most researchers. PEFT has emerged as solution for this challenge, ^3,15^ at it involves training only a small subset of parameters, aiming to match the performance of full fine-tuning with far less compute and memory. As these method update only a small number of additional parameters or update a subset of the pretrained parameters, they preserve the knowledge captured by the LLM model while adapting it to the targeted task. ^16^

The Low-Rank Adaptation (LoRA) is a popularly used PEFT method that adds two trainable low-rank decomposition matrices for weight update. During training, the weights of LLMs are frozen, and only trainable matrices are fine-tuned. During inference, the LoRA weights are merged with the original weight matrix of the LLMs without increasing the inference time. ^17^ Various improved derivates of the LoRA methods had been proposed adding different techniques like dynamically adjusting the rank of LoRA, to improve inference. ^3^ The quantized variant of LoRA – QLoRA, attempts to further reduce the compute and memory foot print by quantizing the model to lower precision ^18^.

This paper investigates whether QLoRA preserves the accuracy gains observed with LoRA, ^19^ while further lowering memory usage and GPU needs. The objective of this work is to make LLM tuning for clinical data-extraction cheaper, faster, and feasible on modest hardware—unlocking broader deployment across health systems.

## Methods

We evaluated whether QLoRA on Llama-3.1-8B-Instruct can approach LoRA’s accuracy on the same model for clinical information extraction while reducing compute. We used Guluzade et. al’s ELMTEX corpus ^19^, for this study. Details are as follows.

### Dataset

The ELMTEX dataset consists of clinical summaries extracted from PubMed Central ^20^. The summaries have been manually annotated with 15 structured information categories (see table 1). We used 13 of the categories, leaving out age and gender. The dataset contains a total of 60,000 clinical cases summaries. We implemented a split with 90% for training and 10% for testing. Only English-language entries were included.

**Table 1.** Distribution of Categories annotations in ELMTEX dataset.

| Categories | Train |  | Test |  |
| --- | --- | --- | --- | --- |
|  | N | % | N | % |
| Medical surgical history | 45523 | 84.3 | 5053 | 84.22 |
| Signs symptoms | 51832 | 95.99 | 5744 | 95.73 |
| Diagnostic techniques procedures | 53395 | 98.88 | 5928 | 98.8 |
| Diagnosis | 53261 | 98.63 | 5915 | 98.58 |
| Laboratory values | 36589 | 67.76 | 4061 | 67.68 |
| Pathology | 39745 | 73.6 | 4390 | 73.17 |
| Pharmacological therapy | 37923 | 70.23 | 4272 | 71.2 |
| Interventional therapy | 39646 | 73.42 | 4334 | 72.23 |
| Patient outcome assessment | 50188 | 92.94 | 5543 | 92.38 |
| Age | 54000 | 100 | 6000 | 100 |
| Gender | 54000 | 100 | 6000 | 100 |
| Family history | 9596 | 17.77 | 1121 | 18.68 |
| Comorbidities | 20280 | 37.56 | 2203 | 36.72 |
| Social history | 4301 | 7.96 | 461 | 7.68 |
| Life style | 12310 | 22.8 | 1309 | 21.82 |

### Model Architecture and Fine-tuning setup

We selected the Llama 3.1 8 billion parameter Instruct Model (Llama-3-1-8b-Instruct) as our base model, given its established effectiveness for following instructions. We fined-tuned the model using LoRA and QLoRA approaches.

Fine-tuning configuration: The base Llama-3-1-8b-Instruct model has 16-bit precision. Hence we quantized the model to 8-bit and 4-bit levels. The configuration utilized dynamic scaling to preserve numerical precision while reducing the memory footprint. We used LORA parameters Rank r = 256, an alpha of 128 and dropout of 0.05. ^19^ To ensure comprehensive adaptation LoRA was applied to all linear layers. Training was conducted for 1 epoch with fused AdamW optimizer, learning rate of 2e-4, batch size of 32 and a maximum sequence length of 5120.

Hardware configuration: We carried out the experiments in Amazon Webservices Cloud using the ec2 g6.12xlarge instance with 700 GB disk drive (gp3), 48 vCPUs and 192 GiB RAM. This machine had 4 NVIDIA L4 Tensor Core GPUs (24 GB VRAM each) and had high network bandwidth (up to ∼40 Gbps). We restricted the training routines to use the minimum require number of GPUs for each of the three experiments, by configuring our code.

Software configuration: All experiments were implemented using python 3.12. Model fine-tuning was conducted with PyTorch, transformers, PEFT, TRL and BitsandBytes libraries. Due to compatibility constrains with 8-bit quantization. In the most recent package release, earlier versions of certain dependencies were adopted to ensure stable execution. The VLLM framework was employed for model deployment. For the evaluation, ROUGE Score, BERTScore, spaCy and sciSpaCy were utilized. The Faiss library was used construct the vector index and retrieve examples for the in-context learning.

Adapter merging: After completing training, we merged the trained QLoRA adapters with base model loaded in bfloat16 precision to ensures high-fidelity of integration.

### Prompting Strategies

We implemented two prompting approaches originally published by Guluzade et al.— Naïve prompt and Advanced prompting with In-Context Learning (prompts are included in Appendix).

Advanced prompting involved a detailed task description with explicit instructions for handling ambiguous cases, and three examples selected using similarity scores.

### Experimental Setup

We first fined-tuned Llama-3.1-8B-Instruct model at full-precision. Next we quantized the model to 8 and 4 bits level, and then fined-tuned the two quantized models. We evaluated these three fined-tuned models, using naïve and advanced prompts.

### Evaluation metrics

We measured the GPU resources required for training each of the three runs. We recorded the number of GPUs and peak GPU RAM used to perform the fine tuning and recorded the time required to complete the training. We also measured the RAM required to load the model for training.

We use the follow metrics for evaluating the performance of the three fine-tuned models.

1. Recall-Oriented Understudy for Gisting Evaluation(ROUGE) Score ^21,22^. We used ROUGE-1, ROUGE-2 and ROUGE-L scores. ROUGE-1 metric captures the overlap of unigrams(single word) between the candidate and reference text. It reflects the model’s ability to preserve basic content units and overall informativeness. ROUGE-2 measures bigram (two-word sequence) overlap, thereby assessing the extent to which the generated summaries capture phrase-level coherence and local word ordering. ROUGE-L is based on the longest common sequence between the candidate and reference summaries. Unlike exact n-gram matching, it accounts for sentence-level structure and fluency, evaluating how well the generated text follows the reference’s overall organization. These scores were calculated using the standard ROUGE library with stemming enabled. (21,22)
2. BERTScore. We also used BERTScore, which will complement n-gram based evaluation. BERTScore assess the semantic similarity between the generated summaries and reference text, in our cases extracted terms with annotations. It leverages contextualized word embeddings to capture deeper semantic alignment. We used microsoft/deberta-xlarge-mnli model, which provides strong general-domain representations optimized for natural language inference. The similarity between candidate and annotated extracted terms was calculated using cosine similarity across token embedding, and we reported F1 Score. ^23^
3. Entity Level Evaluation. Entity-level evaluation is implemented using SciSpacy’s ‘en_core_sci_scibert’ model for medical named entity recognition (NER). Extracted entities were normalized to UMLS Concept Unique Identifiers (CUIs) using SciSpaCY EntityLinker, ensuing standardization of clinical concepts. Evaluation was performed under a strict matching criterion, requiring both entity boundaries and normalized CUIs to match exactly. Metrics included precision, recall and the F1 Score. ^19^

## Results

We developed three fine-tuned models using Llama-3.1-8B-Instruct as the base model. First model was developed by fine-tuning at full-precision (LoRA) and the next two models were developed by fine-tuning after quantizing the base model at 8 and 4 bits precision respectively. The resulting models were evaluated for naïve and advanced prompts, using the following metrics – ROUGE, BERTscore and UMLS entity level precision, recall and F1.

Figure 1 summarizes the results of the evaluation as a heatmap.(see appendix for detailed report) Naïve prompting on LoRA fine-tuned model achieved the highest overall performance across all metrics (B-F1 = 0.82, R1 = 0.79, R2 = 0.69, RL = 0.77, and E-F1 – 0.78)— a significant (10-20 point) improvement over naïve or advanced prompts on the base model. In contrast, the performance of naïve-prompt with QLoRA was only (1-4 pts) lower that LoRA. Advanced prompts performed worse that naïve prompts on both LoRA and QLoRA tuned models. The computational resources used for the model training are shown in table 2.

**Figure 1:**
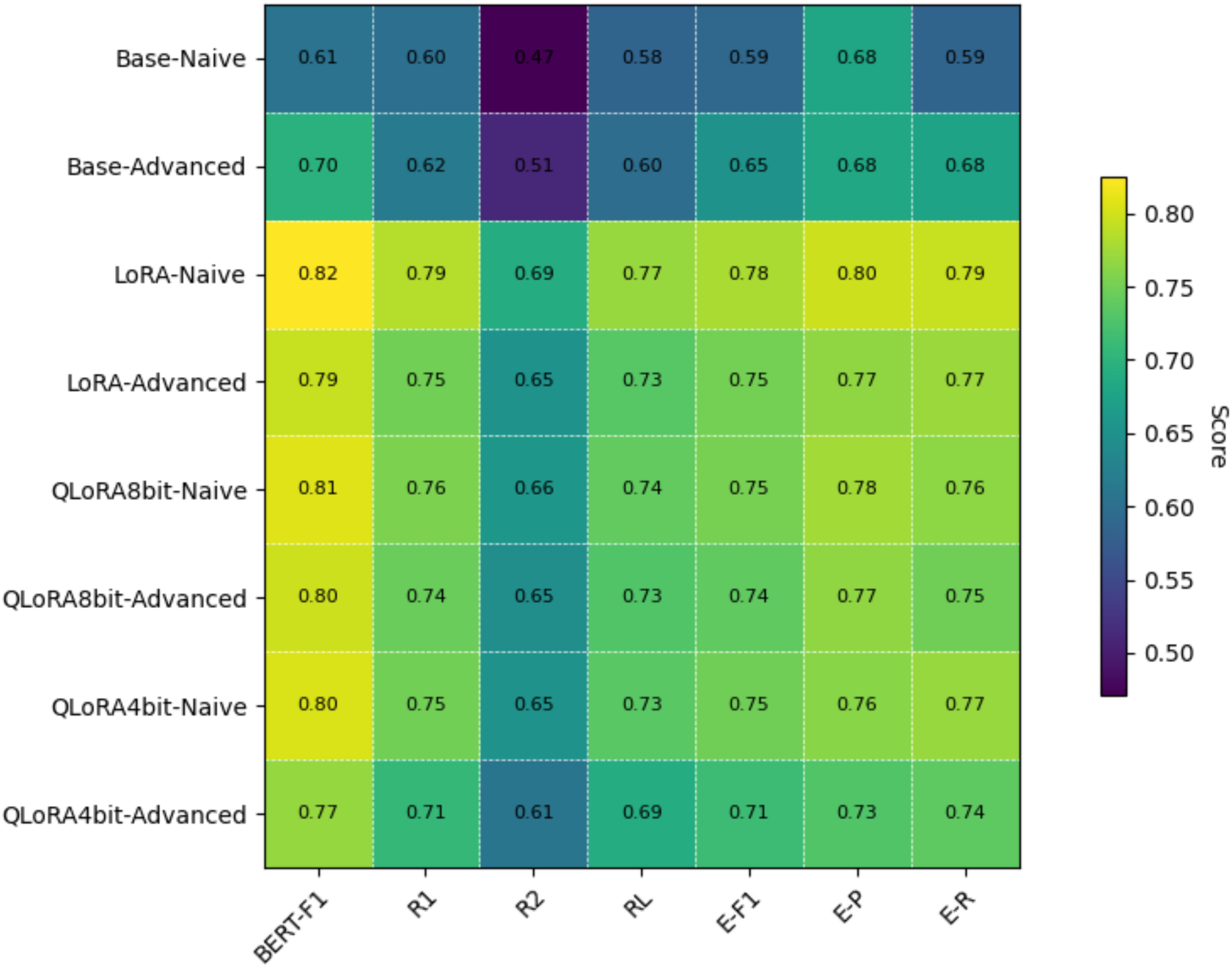
ROUGE, Bert Score and Entity level scores for base model, LoRA fine-tuned, and QLoRA. BF1: BERT-F1, R1: ROUGE-1, R2: ROUGE-2, RL: ROUGE-L, E-F1: Entity F1, E-P: Entity Precision, E-R : Entity Recall

**Table 2.** Computational resources and time needed for the fine-tuning approaches.

|  |  |  |  |  |
| --- | --- | --- | --- | --- |
| Experiment | min # of GPUs | Total GPU RAM use<br>(after Training setup) | Total Peak GPU RAM Use | Time required |
| LoRA | 4 | 14.95 | 35.05 | 28.99 |
| QLoRA_8bit | 3 | 13.6 | 37.03 | 40.18 |
| QLoRA_4bit | 2 | 13.03 | 23.85 | 42.73 |

## Discussion

Fine tuning of LLMs (full-model training) is constrained by the substantial compute and memory demands of conventional methods. As an alternative approach PEFT methods like LoRA have been developed that achieve fine-tuning using less computational resources. PEFT methods work by updating only a small subset of weights, which sharply reduces the computational footprint while preserving near–full-tuning performance. Recent work by Guluzade et al. (18) has shown that fine-tuning LlaMA-3.1.8B-instruct model using LoRA successfully outperforms base models for clinical data extraction. In this work we investigated if further reducing the computational footprint by using quantized models (QLoRA), preserves the accuracy gains observed with PEFT of non-quantized models (LoRA). We used the ELMTEX corpus published by Guluzade etl al. (18), and fine-tuned Llama-3.1-8B at 8-bit and 4-bit quantization.

Our results show that fine-tuning markedly improved performance across all metrics indicating that PEFT methods can lead to significant improvement over prompting strategies. In our study, Naïve prompting on LoRA fine-tuned model achieved the highest overall performance across all metrics. There was a accuracy gain of 10-20 points compared to the base model. Even the quantized models had an improvement of 8-14 pts over the base model.

The performance of naïve-prompt with QLoRA was only (2-4 pts) lower that LoRA, which indicates that QLoRA preserves most of the accuracy gains (10-20 pt) obtained with LoRA. However QLoRA requires much lower compute and memory resources.

LoRA fine-tuning need 4 GPUs, while QLoRA could be completed in 3 or 2 GPUs for 8-bit and 4-bit models respectively. Additionally the QLoRA models required 10-13% less memory for loading the model for training. Notably 4-bit QLoRA needed only 68% of peak GPU RAM as compared to LoRA. However 8-bit QLoRA required 106% of peak GPU RAM as compared to LoRA. The latter is possibly due to lower optimization support for 8-bit optimization.

However QLoRA needed 40-42 hours for training as compared to only 29 hours for QLoRA—It was 28%-32% slower. This is possibly due to overhead for dequantizing the weights as the back propagation is still performed in 16 bit precision, and that the training libraries are not matured to optimize 8/4 bit precision^24^.

Our results also show that advanced prompts performed worse than naïve prompts on both LoRA and QLoRA tuned models. This is likely because the fine-tuning was performed using naïve-prompts.

Overall our study validates several important trends in the application of parameter-efficient fine-tuning for domain-specific extraction. The results confirm that prompting alone provides limited but measurable improvements in baseline models. As seen with advanced prompting strategy, gains in both ROUGE and entity-level f1 indicate that carefully designed prompts can guide models towards more accurate outputs. However, these improvements are relatively modest compared to the fine-tuned model suggesting that prompting cannot fully compensate for the lack of domain adaptation.

Advancing PEFT methods is critical to enabling more teams using modest hardware to tune LLMs for specific clinical tasks. Resource constraints are common in hospital IT infrastructure and our results show that 4-bit QLoRA can offer a promising approach that can be practically implemented on modest hardware.

### Limitations

A major limitation of our study is that we used a single model architecture (LLaMA-3.1-8B), which limits its generalizability across LLMs or parameter sizes. Additionally, the dataset used is clinical case reports which might not have complexity of real-world clinical notes. Future work should explore QLoRA’s performance across a broader range of model architectures and sizes utilizing unstructured clinical datasets.

## Conclusion

This study demonstrates a comparative analysis of QLoRA fine-tuning and hard-prompting techniques for structured data extraction from clinical notes. Results show that QLoRA preserves most of the accuracy gains obtained with LoRA, and requires much lower compute and memory resources. By reducing the memory foot-print it offers a practical solution for making advanced clinical NLP more accessible for accurate data extraction in resource-constrained environments. Future work should test QLoRA across diverse architectures and sizes and on clinical corpora representative of real-world practice.

## Supplementary files

1. Supporting Tables and Figures
2. Prompts

## Data availability

The source code we developed for this study is published at Github-link https://github.com/dschc/qlora-vs-lora-clinical-SDE.git The data used in this study was made publicly available by Guluzade et. al. at https://zenodo.org/records/14793810.

## Acknowledgement

We are thankful to Guluzade et. al. for publishing their data-set and source-code in the public domain, that enabled this study. This project was supported by grants from NHLBI (R01-HL151643) and Amazon Web services.

## Annex 1: Figures and Tables

**Figure S1:**
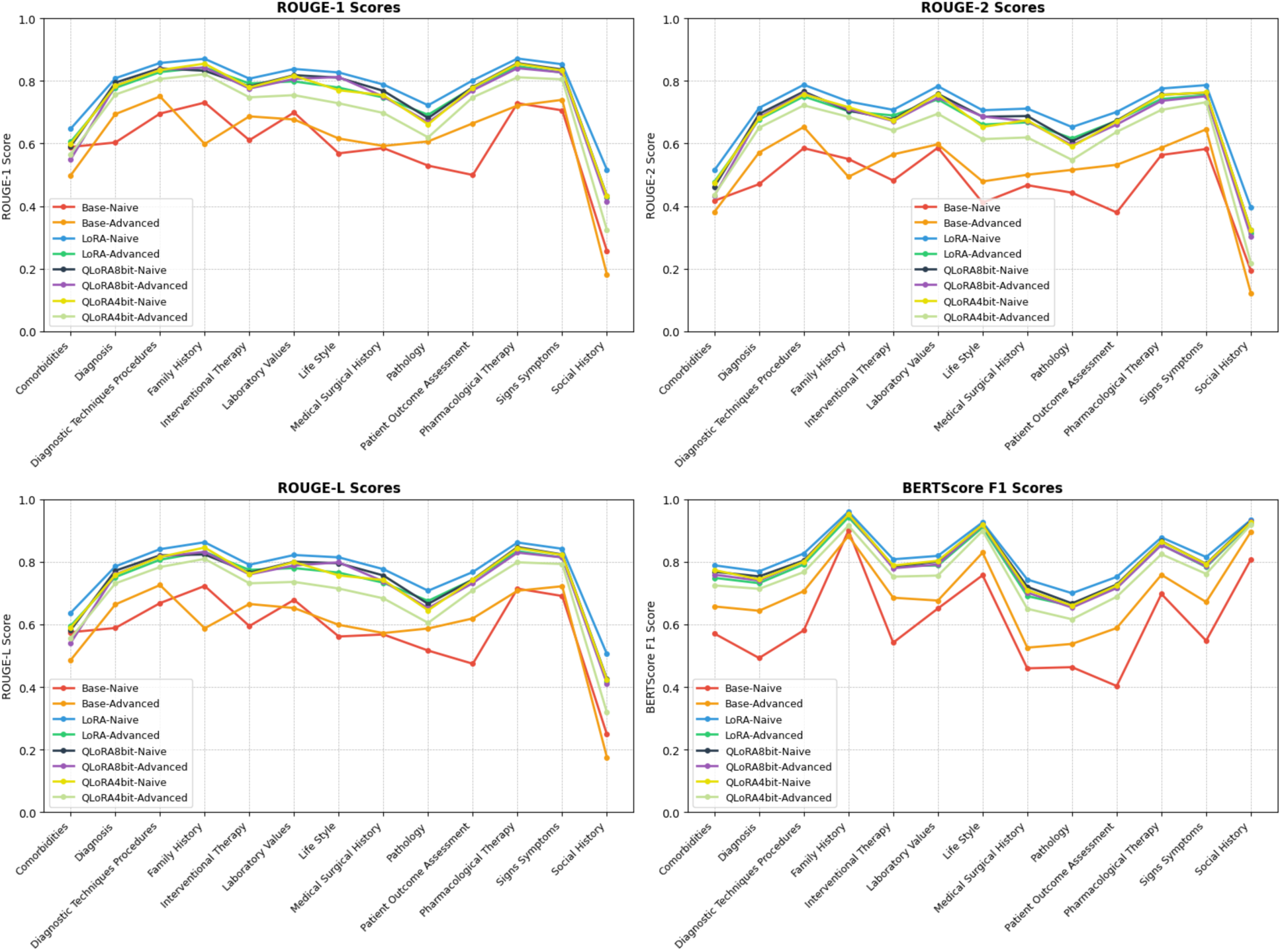
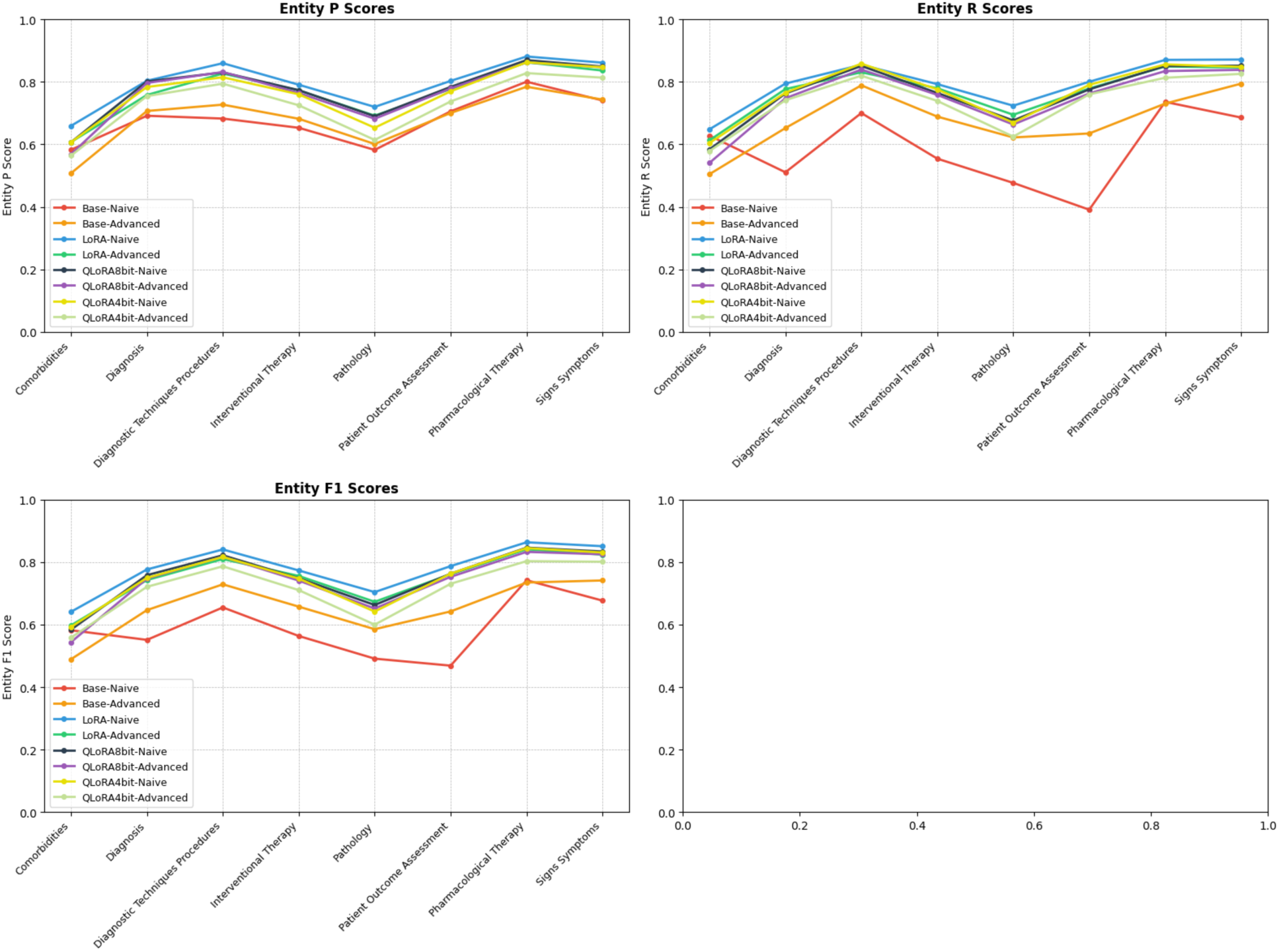
Model Performance across structured data extraction categories (ROUGE, BERT and Entity level Precession(P), Recall(R) and F1 score).

**Table S1:** Comparison of naïve prompt results of 4bit-QLoRA fine-tuned model trained for 1 epoch vs 3 epoch.

|  |  |  |  |  |  |  |  |
| --- | --- | --- | --- | --- | --- | --- | --- |
| Model | B-F1 | R1 | R2 | RL | E-F1 | E-P | E-R |
| QLoRA4bit 3epoch | 0.82 | 0.79 | 0.69 | 0.77 | 0.78 | 0.79 | 0.79 |
| QLoRA4bit 1epoch | 0.80 | 0.75 | 0.65 | 0.73 | 0.79 | 0.76 | 0.77 |

## Annex 2: Prompts

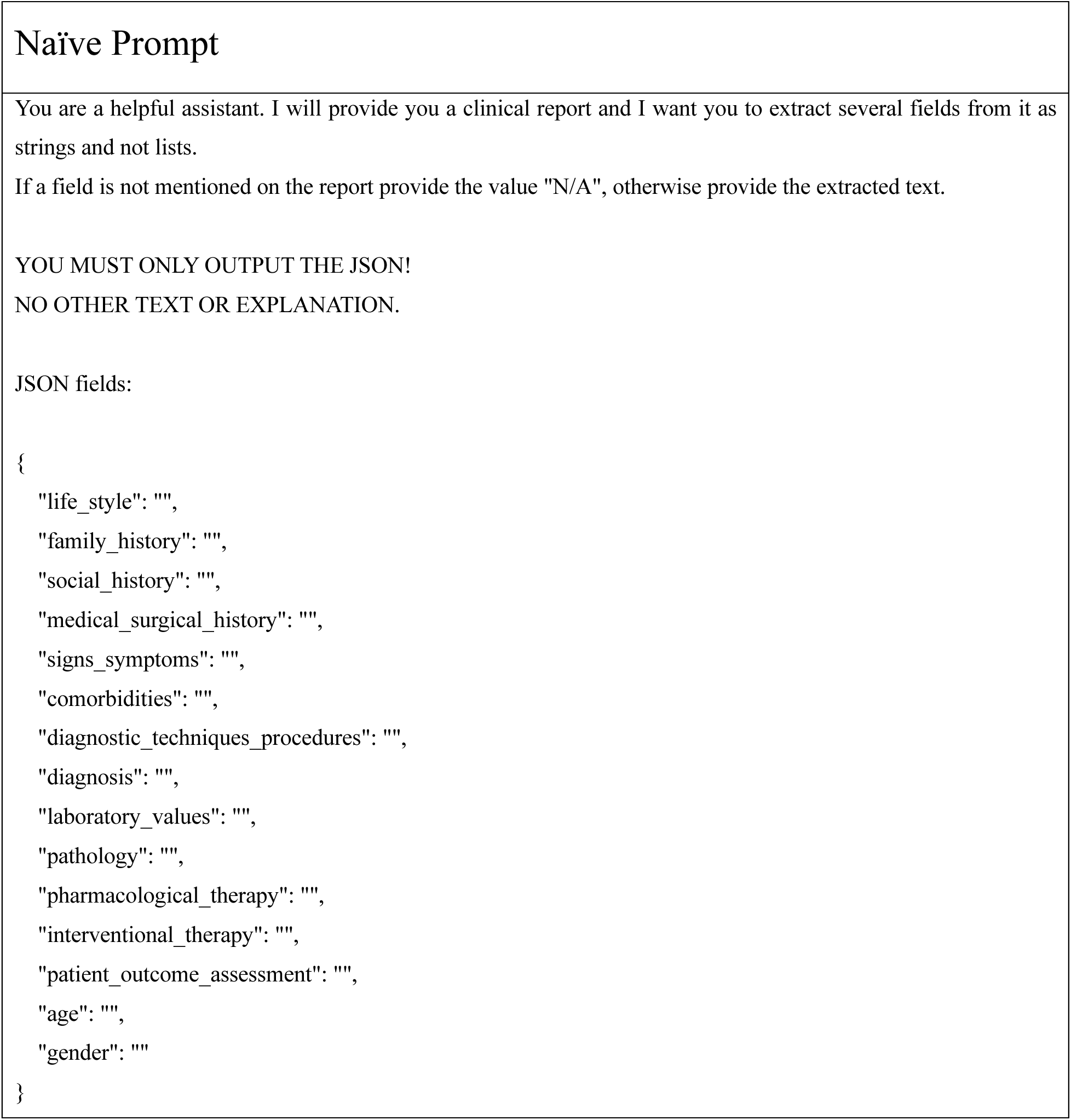

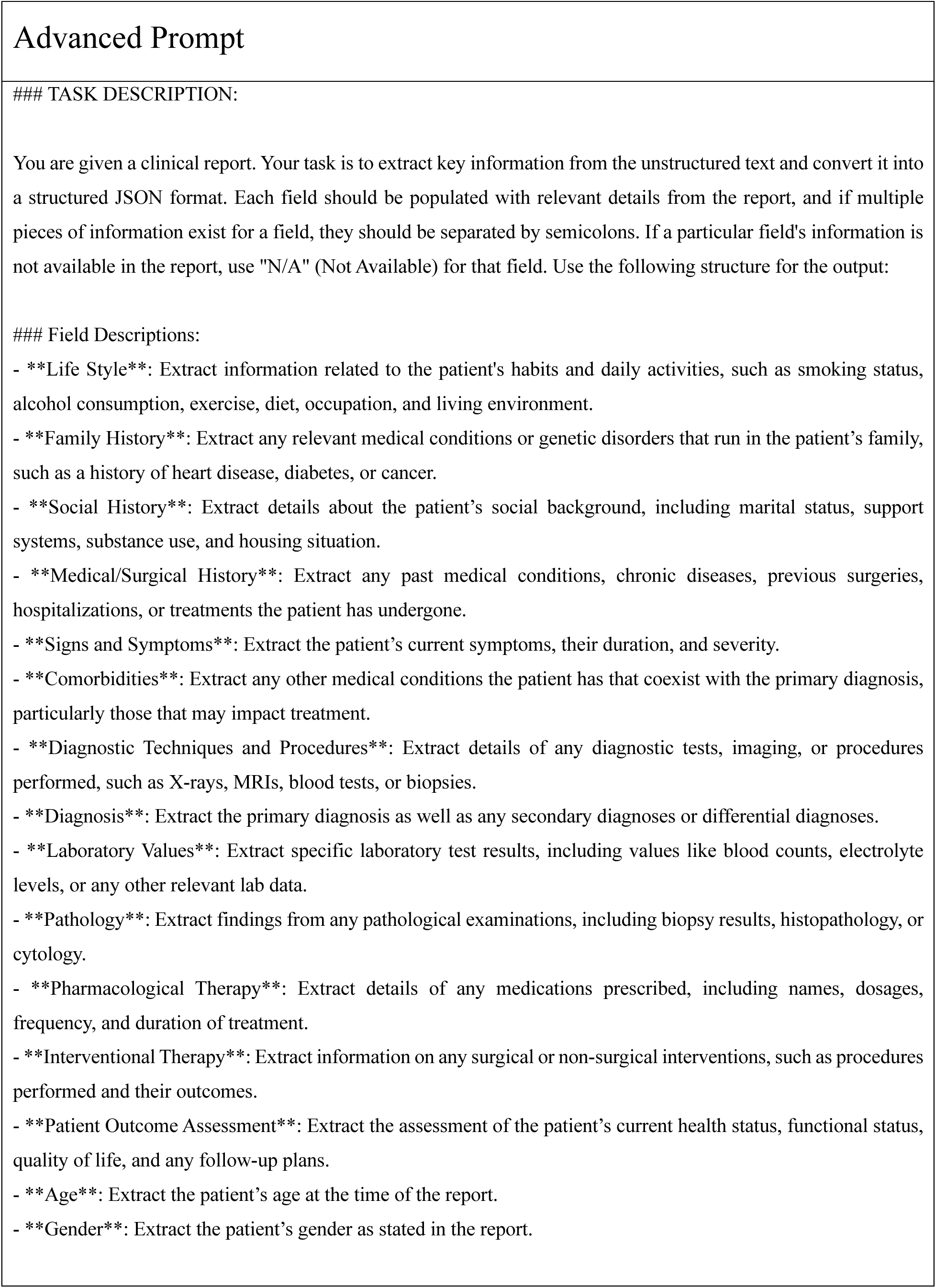
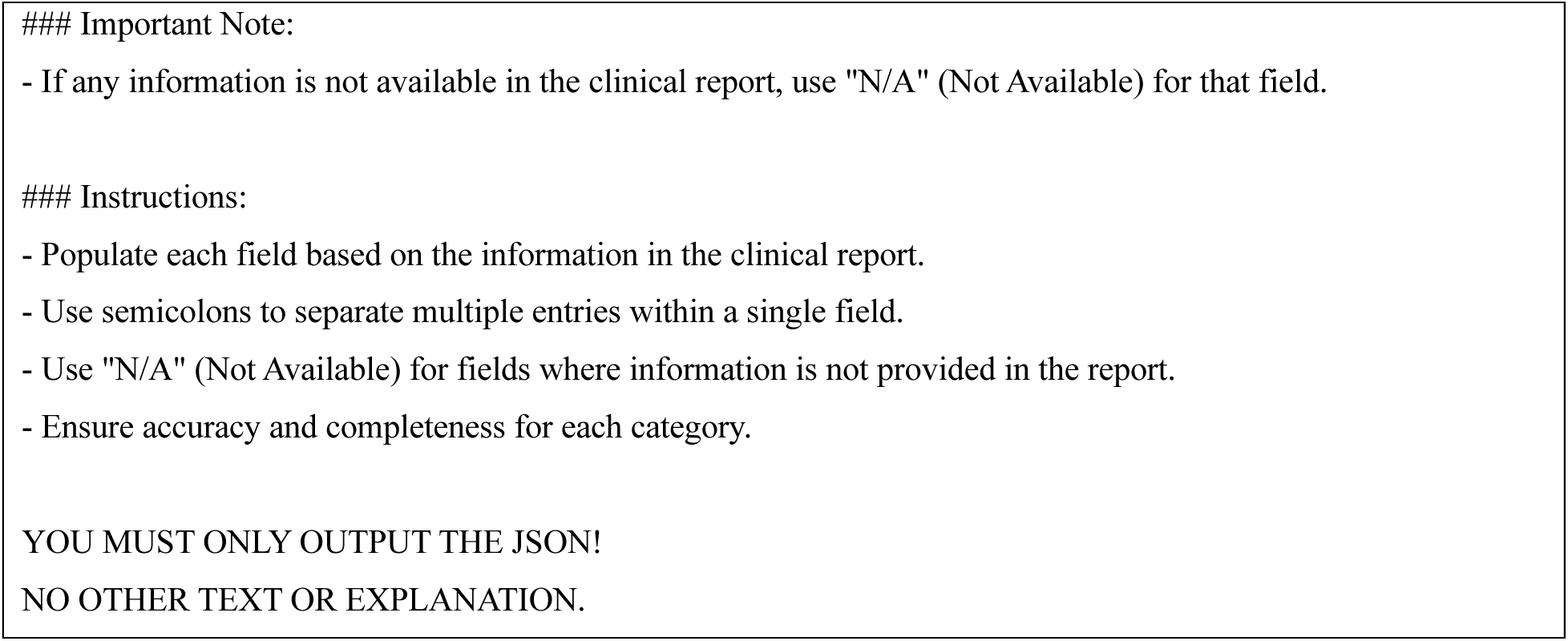

